# Reduced POLD3 Expression Induces DNA Damage and Telomere Shortening in Colorectal Cancer Cells: A Potential Mediator of Colorectal Cancer Risk Variants

**DOI:** 10.64898/2025.12.19.25342655

**Authors:** Eloise Clarkson, Nagore De LeoN, Sarah Aldulaimi, Ian Tomlinson, Terry Roberts, Annabelle Lewis

**Affiliations:** Centre for Genome Engineering and Maintenance, Department of Biosciences, College of Health and Life Sciences, Brunel University London, Uxbridge, UB8 3PH, UK; Centre for Genomics and Child Health, Blizard Institute, Queen Mary University of London, E1 2AT, UK; Department of Oncology, University of Oxford, Old Road Campus Building, Roosevelt Drive, Oxford, OX3 7DQ

## Abstract

DNA Polymerase Delta (Pol δ) acts in DNA synthesis and in DNA repair, both important factors in the proliferative nature, longevity, and mutability of cancer cells. The chromosomal region containing *POLD3*, the third subunit of Pol δ, has recently been identified by genome wide-association studies as containing single nucleotide polymorphisms (SNPs) that increase the risk of colorectal cancer (CRC).

To explore the effects of changing *POLD3* levels that may underly these statistical associations in CRC, we first carried out survival analyses in the Cancer Genome Atlas colon adenocarcinoma cohort. Cox proportional hazards modelling showed higher POLD3 expression to be associated with moderately improved overall survival in the full cohort. This association with survival was clearest in the microsatellite stable (MSS) subgroup.

In order to investigate the mechanisms by which lower POLD3 expression may be associated with poorer outcome, we generated CRC cell line models with reduced POLD3 expression levels. Knockdown of POLD3 resulted in reduced proliferative ability and stalling of the cell cycle in S phase, with some apoptosis of CRC cells. Importantly, POLD3 knockdown cells exhibited DNA damage, manifested by double-stranded breaks (DSBs), and upregulation of DSB repair pathways. Reduced POLD3 also correlated with shortened telomeres with disruption of, specifically, the ALT telomeric maintenance pathway.

Taken together these observations suggest that lower POLD3 expression can increase genetic instability in CRC cells in the form of DSBs and shortened telomeres and that this might underly the functional effect of CRC risk SNPs in the POLD3 locus in 11q13.4.

**Simple Summary:** DNA variants associated with an increased risk of colorectal cancer (CRC) are found near the gene POLD3. This study investigated the role of POLD3, a DNA replication and repair protein, in CRC. Analysis of patient data suggested that lower POLD3 expression was associated with reduced overall survival, particularly in patients with microsatellite stable tumours. To understand the biological basis of this association, CRC cell lines with reduced POLD3 expression were generated. Lower POLD3 levels slowed cell growth, stalled DNA replication, and increased cell death. Importantly though, reduced POLD3 expression led to greater DNA damage, including double-strand breaks, and activated DNA repair pathways. Cells with lower POLD3 also had shorter telomeres, probably due to disruption of the alternative lengthening of telomeres (ALT) maintenance pathway. These findings suggest that reduced POLD3 expression promotes genetic instability in CRC cells, which may contribute to poorer clinical outcomes, and provide a possible functional explanation for the risk variants in the POLD3 genomic region.

## Introduction

Cancer progression is frequently promoted by genomic instability in the form of increased mutation rate or larger scale chromosomal gains, losses and rearrangements. Such instability has multiple origins including disrupted DNA replication and repair pathways, ineffective cell cycle checkpoints and telomere dysfunction.

Colorectal cancer (CRC) is one of the most prevalent and deadly cancers globally, and accounts for 10% of all new cancer cases each year (1). Genomic instability is a hallmark of CRC, and CRC can be classified by different forms of genome or (epi)genetic instability, including chromosomal instability (CIN), microsatellite instability (MSI) caused by faults in the mismatch repair (MMR) system, and high single nucleotide mutation rates (hypermutation-ultra mutation) (2). Several CRC predisposition syndromes such as LYNCH, MAP and PPAP result from heritable mutations in DNA repair pathways (3–5) , but somatic mutations and epimutations in DNA repair and replication pathways are also common. Mutations in the major DNA polymerases, such as POLE and POLD1, have been shown to cause decreased proofreading, genome instability, and cancer. One recurrently mutated DNA replication and proof-reading enzyme complex is Pol δ.

Recent genome-wide association studies (GWAS) have identified a CRC risk locus, 11q13.4, which contains the POLD3 gene, the third subunit of Pol δ (6, 7). The first CRC risk SNP in the region to be identified was a POLD3 intronic SNP rs3824999 from a GWAS meta-analysis, with imputation indicating a further, stronger association with rs72977282 mapping to the POLD3 region (6). There is, as yet, no functional evidence confirming that these SNPs are causal, or any conclusive data as to how they may modify gene expression on chromosome 11q13.4. However, POLD3 is an excellent candidate as a target gene of one or more of these SNPs due to its role in DNA replication and repair and the characterised roles of other DNA polymerase subunits in CRC.

The polymerase Delta complex (Pol δ) has a role in both synthesis of DNA and its repair, making it a clear functional target for maintenance of DNA integrity. Pol δ is a heterodimer comprising subunits POLD1, POLD2, POLD3, and POLD4. During DNA replication, Pol δ is responsible for lagging strand DNA synthesis, as well as repair of DNA double-strand breaks (DSBs) via homologous recombination (HR). Pol δ works as a gap-filling polymerase, which is often a site for DNA fragility during replication (8).

POLD3 has dual roles in the Pol δ complex, first as a stabiliser of the POLD1-POLD2 interaction, and second as a facilitator to the binding of the POLD complex to PCNA through a C-terminal PIP box. POLD3 also exhibits 3’ to 5’ exonuclease activity, which increases the processivity of DNA synthesis during replication. Removal or misfunction of the POLD3 subunit prevents PCNA binding to Pol δ, and therefore ineffective DNA replication (9,10).

In addition, Pol δ has roles in multiple repair pathways (9). These include the DNA mismatch and base-excision repair pathways that are defective in the hereditary CRC susceptibility disorders, Lynch syndrome and MAP respectively (10, 11). POLD3, in particular, is important for break – induced replication repair (BIR) and also alternative telomerase independent telomere maintenance (ALT) pathways (12, 13). In contrast to telomere maintenance in tumour cells carried out through telomerase reactivation via re-expression of the TERT gene (14), the ALT mechanism of telomere maintenance is less well understood. It is thought to be activated by R-loops, G-quadruplexes, and DNA single-strand breaks causing the collapse of replication forks. These induce SUMOylation of telomere proteins, triggering recruitment of ALT-associated PML bodies, which initiate DNA repair by break-induced repair (BIR) mechanisms. BIR at telomere ends is mediated by POLD3/POLD4 and the production of C-circles at telomere ends (15).

Recent studies have shown that POLD3 loss in mouse embryonic stem cells results in rapid telomere shortening, chromosomal abnormality and aneuploidy, and increased number of chromosome or chromatid breaks (16). The known cross-over between the BIR and ALT pathways supports the fact that POLD3 is a probably central player in both mechanisms (12, 17). Its disruption or loss in cancer could cause DNA strand breaks in the main body of chromosomes and also at the telomeres, both mechanisms of genomic instability.

Given the strong genetic association of POLD3 locus variants with CRC and its known role in replication and repair, we wished to further explore the potential of POLD3 as a target for these variants and its role in replication and genomic instability in CRC cells .To demonstrate the functional effects of reduced POLD3 on CRC cells, we have investigated the proliferative, cell cycle, and DNA repair changes upon POLD3 knockdown. Furthermore, we have shown the effect of POLD3 knockdown on telomere maintenance, a crucial component of cancer cell genomic stability. Finally we examined CRC patient data to determine the effect of POLD3 expression levels on survival.

## Results

### POLD3 expression is associated with overall survival in colon adenocarcinoma patients

We evaluated POLD3 expression for its clinical relevance in The Cancer Genome Atlas (TCGA) colon adenocarcinoma (COAD) cohort. POLD3 expression differed according to molecular subtype, being higher in MSI tumours than in MSS cases and enriched in CMS1/CMS3 relative to CMS2/CMS4 (Figure 1A and B, Supplementary Table S1). In addition, POLD3 expression varied significantly across pathological stage, with post hoc analyses indicating that this effect was driven predominantly by lower expression in stage IV tumours (Supplementary Figure S1, Supplementary Table S1). These findings indicate that POLD3 levels are shaped by both molecular and clinical tumour context.

**Figure 1:**
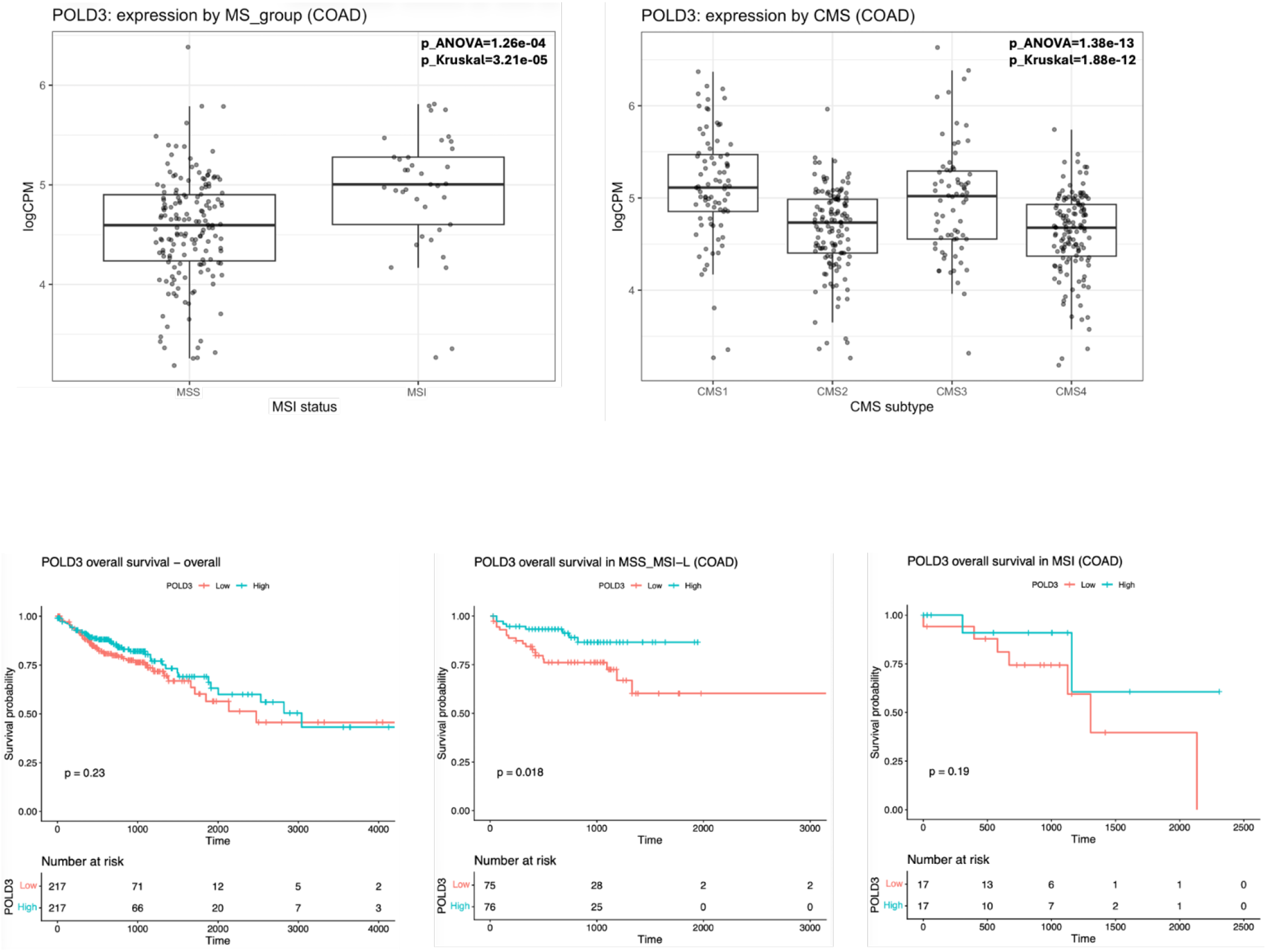
POLD3 expression analysis across molecular subtypes in TCGA colon adenocarcinoma (COAD) and effects on patient survival. A) Left panel. POLD3 expression in MSS and MSI tumours. Right panel. POLD3 expression across consensus molecular subtypes (CMS1-CMS4). Expression values are shown as logCPM from RNA-seq data. Boxplots represent the median and interquartile range, with individual tumours overlaid. Statistical comparisons were performed as described in Methods section. **B)** Kaplan-Meier analyses of overall survival in TCGA COAD stratified by median POLD3 expression in the full cohort and within microsatellite subgroups. Left, overall COAD cohort plogRank=0.23, n=434, 95 events; centre, MSS subgroup plogRank=0.018, n=151, 27 events; right, MSI subgroup plogRank=0.19, n=34, 9 events. Across analyses, lower POLD3 expression showed an overall tendency towards poorer survival, with the clearest separation observed in the MSS subgroup.

We next used Kaplan-Meier analysis to investigate the association of POLD3 expression levels with patient outcome. However, this median-based stratification did not show a statistically significant difference in the full cohort although the direction of effect was consistent with poorer outcome in tumours with lower POLD3 expression (Figure 1C, Supplementary Table S2). Consistent with this pattern, Cox proportional hazards modelling using continuous expression values identified higher POLD3 expression as significantly associated with improved overall survival (HR = 0.780, 95% CI 0.637-0.954, p = 0.016; n = 434, 95 events, Supplementary Table S3). Stratified analyses showed the clearest survival separation in the MSS subgroup (Kaplan-Meier, p=0.018 n=151, Cox HR = 0.722, 95% CI 0.502-1.039, p = 0.080; n = 151, 27 events Figure 1C, Supplementary Table S2 and 3) whereas no robust difference was observed in MSI tumours; however, interaction testing did not support a strong differential effect by microsatellite group. Exploratory survival analyses stratified by CMS subtype are provided in the Supplementary material (Supplementary Figure S2, Supplementary Table 3) showing stronger effects of POLD3 expression on the CMS1 and CMS3 subtypes.

### POLD3 knockdown reduces proliferation and inhibits cell-cycle progression

Our survival analyses provide suggestive evidence that low POLD3 expression is associated with poorer outcome in CRC patients. We wished to explore this further by investigating the effect of reducing POLD3 on CRC cells. To test whether modest decreases in POLD3 dosage are sufficient to perturb epithelial cancer cell behaviour, we modelled reduced POLD3 expression in colorectal cancer cell lines.

To this end, we transduced 2 well-characterised CRC cell lines, CACO2 and RKO, with virally packaged doxycycline-inducible shRNAs targeting the POLD3 gene. To control for off-target effects, 2 shRNAs targeting different areas of the POLD3 gene were generated, which were transduced to create two separate CACO2 and two separate RKO cell lines, termed POLD3 shRNA1 and POLD3 shRNA2. Doxycycline (Dox) was administered to cells lines at low (0.1 μg/ml), medium (1 μg/ml) and high levels (10 μg/ml) to analyse the effects of increasing POLD3 depletion. qPCR and western blotting confirmed mRNA and protein reduction which varied in a dose dependent manner (Figure 2A and Supplementary Figure 3A).

**Figure 2:**
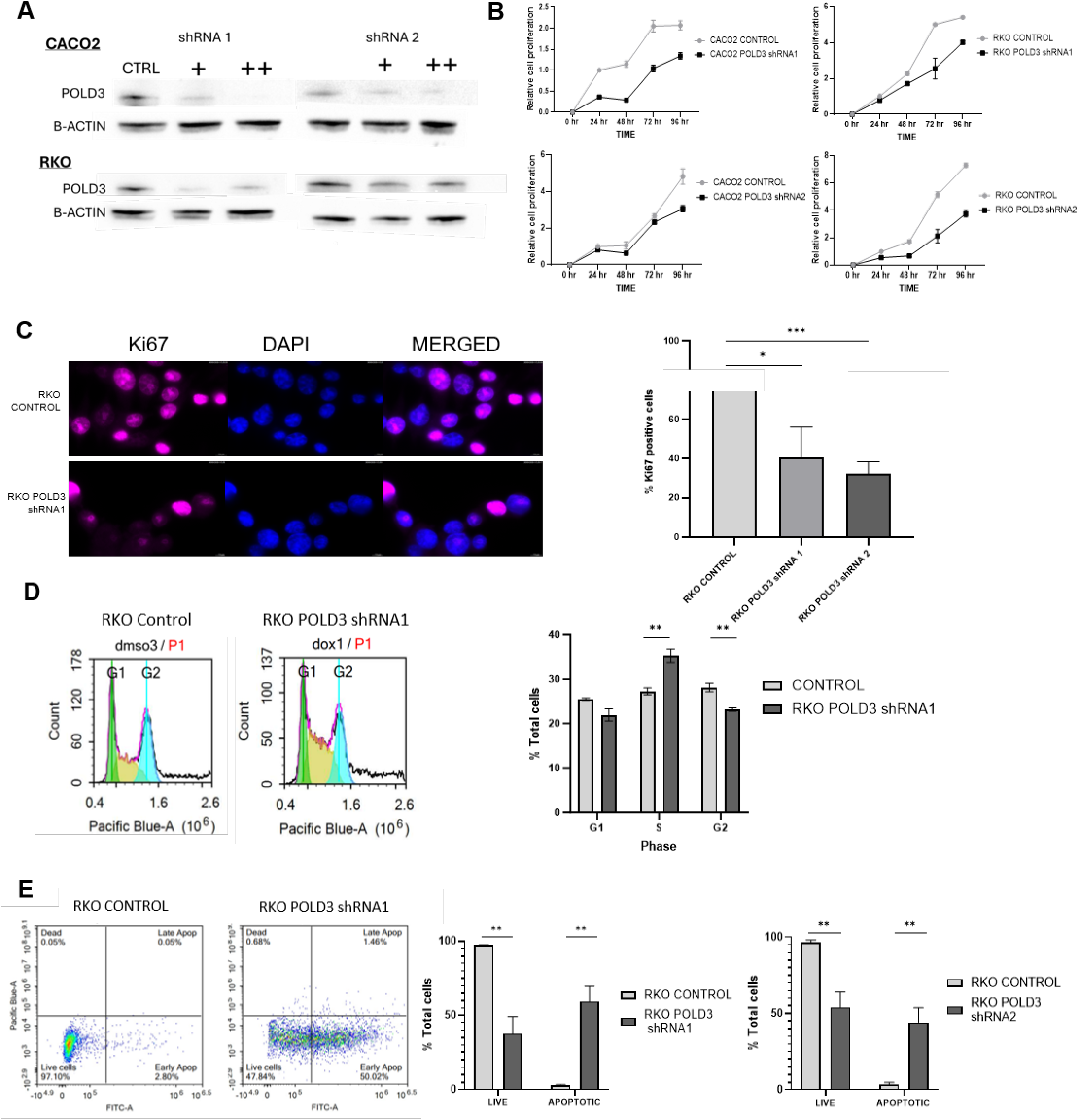
POLD3 knockdown restricts cancer cell growth due to cell cycle changes and increased apoptosis. A) Western blotting to measure POLD3 protein levels in cell lines with lentiviral transduction of shRNA1 and shRNA2 POLD3 knockdown constructs. Cells were treated with 0 (DMSO only), 1 μg/ml or 10 μg/ml doxycycline for 48 hours to induce shRNA expression and POLD3 knockdown. B) Cellular proliferation analysis, CACO2 and RKO POLD3 shRNA 1 and 2 knockdown cells were treated with 0 or 10 μg/ml doxycycline for 48 hours to induce shRNA expression. In both CACO2, and RKO cell lines, proliferation was significantly decreased after shRNA 1 and shRNA2 induction, T-test; n= 3, P<0.0001, P<0.001. C) Upper panel: Representative images of Ki-67 immunofluorescence staining of RKO POLD3 shRNA1 knockdown cells treated with 0 or 10 μg/ml doxycycline for 48 hours to induce shRNA expression. Lower panel: quantification of the percentage of Ki-67+ cells with immunofluorescence staining in RKO POLD3 shRNA 1 and shRNA 2 knockdown cells induced with 10 μg/ml doxycycline. Students T-test: POLD3 shRNA1 P<0.05, POLD3 shRNA2 P<0.001. D) Upper panel: Histograms of flow cytometry analysis of cell cycle proportions in RKO POLD3 shRNA1 knockdown cells induced with 10 μg/ml doxycycline. Lower panel: Quantification of cell cycle stage of RKO POLD3 shRNA 1 induced with 10 μg/ml doxycycline. T-test. shRNA 1: G1 P=NS, S P<0.01, G2P<0.01. E) Left panel: Scatter plots of apoptotic cells in RKO POLD3 shRNA 1 and shRNA 2 knockdown cells induced with 10 μg/ml doxycycline, stained with apoptotic marker Annexin V. Right panel: Quantification of apoptotic cells marked by Annexin V staining via flow cytometry in RKO POLD3 shRNA 1 and shRNA 2 knockdown cells induced with 10 μg/ml doxycycline. T-test: shRNA1 live cells P<0.01 apoptotic cells P<0.01, shRNA2 live cells P<0.01 apoptotic cells P<0.01. Error bars given as ± SEM in all panels.

POLD3 knockdown was efficient in both cell lines using shRNA1, with 10 μg/ml doxycycline treatment of CACO2 POLD3 shRNA1 cell lines showing a significant reduction of POLD3 mRNA expression to 35% of normal levels (P<0.01) and RKO to 50 % (P<0.01). shRNA2 also showed significant reduction of POLD3 expression in CACO2 cells at 10 μg/ml to 60% (P<0.005), however this reduction was less than that of shRNA 1. In RKO cells, shRNA 2 showed a greater reduction of POLD3 expression than shRNA 1 at 30%, with 0.1 μg/ml Dox (P<0.01) and 10 μg/ml Dox (P<0.01). At the protein level, a substantial reduction of POLD3 was seen in both cell lines with shRNA1 performing better.

Proliferation of cancer cells inherently relies on the use of DNA polymerases to synthesise DNA in S-phase of the cell cycle. Therefore, we confirmed that POLD3 knockdown inhibited proliferation in our cell lines. Both CACO2 and RKO showed a reduction in cellular proliferation after 72hrs treatment with 10 μg/ml doxycycline to induce shRNA 1 expression (P<0.0001 and P<0.05 respectively). RKO cells also showed a greater decrease in proliferation under shRNA 2 induction, at all induction levels (P<0.001, P<0.001, P<0.001) (Figure 2B). This was supported by a reduction ki67 positive cells in our POLD3 knockdown population. Representative images and graphs in Figure 2C show immunofluorescence staining of Ki67 was present in around twice the number of control cells compared to that of in our POLD3 knockdown cell lines (shRNA1, P<0.05; shRNA2, P<0.0001).

Next, we analysed the effects of POLD3 on cell cycle progression. Synchronised knockdown cells were generated by arrest in G1 phase through serum withdrawal, and release through serum addition, in parallel with POLD3 shRNA induction. As seen in Figure 2D, shRNA1 significantly increased the proportion of RKO cells stalled in S phase (P<0.005) while reduced the proportion of cells in G1 and G2. Similar results were seen for shRNA2 (Supplementary Figure 3B). This tendency to stall in S phase signifies a reduction of DNA replication ability and/or checkpoint activation during POLD3 knockdown.

As seen in figure 2E, flow cytometry staining with Annexin V revealed that POLD3 knockdown increased the proportion of cells that had begun to enter apoptosis. This was found in both RKO shRNA1 (P<0.01) and shRNA2 (P<0.01) cell lines. This data collectively suggests that POLD3 knockdown causes a reduction in cell proliferation due to S-phase stalling, and increases cell apoptosis.

### POLD3 knockdown induces Double-stranded breaks and DNA damage in CRC cells

The other major function of Pol δ is DNA repair, and POLD3 in particular has been extensively shown to be required for break-induced replication (BIR) (16, 18). Therefore, DNA strand breaks (SBs) both single and double were analysed through an alkaline-comet assay. CACO2 and RKO cells were treated with doxycyline to induce POLD3 shRNA1 mediated knockdown for 2 weeks to induce SBs. As seen in Figure 3A, CACO2 knockdown cells showed increased comet “tail” length after gel electrophoresis, indicating increased SBs due to POLD3 depletion. Quantification of tail length from both CACO2 and RKO knockdown cells showed significantly increased tail length in both (P<0.0001 and P<0.01 respectively).

**Figure 3:**
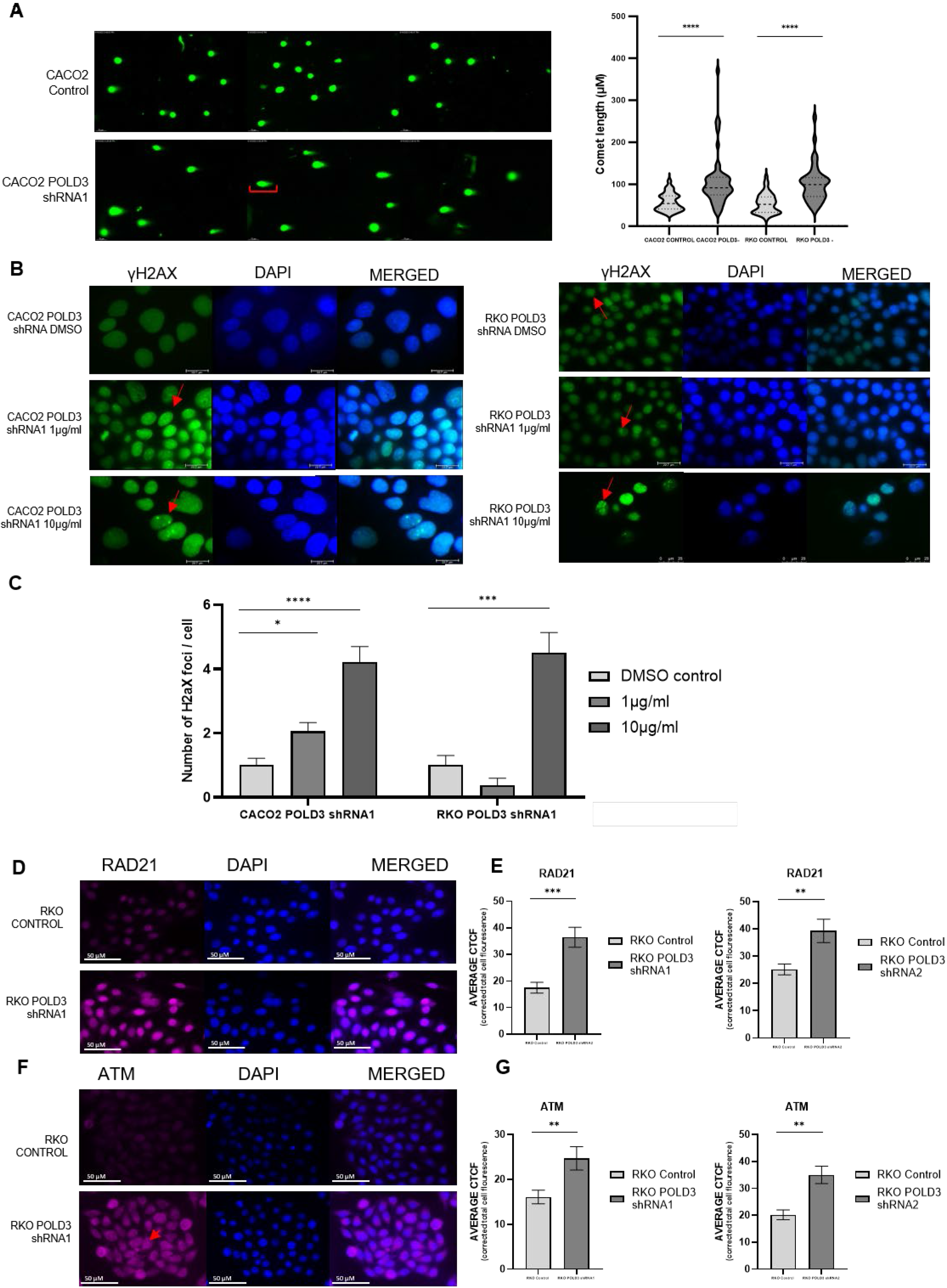
POLD3 knockdown increases DNA damage in CRC cells A) Left panel: Representative images of comet assay of CACO2 and RKO POLD3 shRNA1 cells treated with 0 or 10 μg/ml doxycycline for 48 hours to induce POLD3 knockdown. Right panel: Quantification of Comet tail length in control and knockdown cells. T-test: CACO2 shRNA1 P<0.0001. RKO shRNA 1T-test P<0.0001. N=3 (50 comet tails measured per replicate). Quantification carried out on Image J. Red arrow depicts comet length. B) Representative images of immunofluorescence detection of γH2AX on CACO2 POLD3 shRNA1 (left) and RKO POLD3 shRNA1 (right) cells treated with 0 (DMSO), 1μg/ml or 10μg/ml doxycycline to induce shRNA expression and POLD3 knockdown (representative images). Cells were incubated for 72hrs before γH2AX detection. C) Quantification of γH2AX foci in CACO2 POLD3 shRNA1 cells treated for 72 hrs. 0 (DMSO), 1μg/ml or 10μg/ml doxycycline to induce shRNA expression and POLD3 knockdown. Cells were incubated for 72hrs before γH2AX detection. CACO2 POLD3 shRNA1 1μg/ml P< 0.05, 10μg/ml P<0.0001; RKO POLD3 shRNA1 10μg/ml P<0.001. N=3. D) Immunofluorescence staining of RAD51 on RKO POLD3 shRNA1 cells treated with 0 (DMSO), 1μg/ml or 10μg/ml doxycycline to induce shRNA expression and POLD3 knockdown. (representative images (left)).E) Immunofluorescence quantified as Corrected Total Cell Fluorescence (CTCF). RKO POLD3 shRNA1 10μg/ml P<0.001; RKO POLD3 shRNA2 10μg/ml P<0.01 N=3 T-test. F) Immunofluorescence staining of ATM on RKO POLD3 shRNA1 cells treated with 0 (DMSO), 1μg/ml or 10μg/ml doxycycline to induce shRNA expression and POLD3 knockdown. (representative images (left)). G) Immunofluorescence quantified as Corrected Total Cell Fluorescence (CTCF). RKO POLD3 shRNA1 10μg/ml P<0.01; RKO POLD3 shRNA2 10μg/ml P<0.01 N=3 T-test N=3 T-test. Error bars given as ± SEM in all panels.

We used the marker γH2AX to demonstrate that this DNA damage included double strand breaks (DSB, Figure 3B). In both CACO2 POLD3 shRNA1 and RKO POLD3 shRNA1 cell lines, γH2AX foci indicate an accumulation of DSBs following POLD3 knockdown. In CACO2 cells, DSBs were increased with decreased levels of POLD3 at 1 μg/ml Dox and 10 μg/ml Dox (P<0.05 and P<0.0001, Figure 3C). In RKO cells, the highest level of POLD3 knockdown also significantly increased the presence of γH2AX foci (P<0.001).

We then assessed whether POLD3 knockdown would lead to an increase in DNA repair pathway signalling due to the formation of DSBs. RAD21 is central mediator of DNA repair via HR and its levels increased during POLD3 knockdown, in dox induced RKO cells with both POLD3 shRNA1 and shRNA2 (P<0.0001, P< 0.01; Figure 3D and E). We also analysed the presence of the transducer protein, ATM. Again, ATM levels were increased during POLD3 knockdown by shRNA1, (P<0.0001, P<0.001; Figure 3 F and G). Together this data indicates that POLD3 is required for successful DNA replication, and knockdown of the POLD3 gene results in DNA damage including the formation of DSBs, and therefore initiation of the DSB repair pathway.

### POLD3 knockdown reduces telomere length in colorectal cancer cells

Telomeric length is often regarded as a paradox in cancer cells with activation of maintenance mechanisms increasing longevity but shortened telomeres increasing genomic instability. The majority of cancer cells maintain telomeres through telomerase reactivation, whereas a small subset of cells (10∼15%) also rely on the alternative lengthening of telomeres (ALT) mechanism (19, 20). However, recent publications have discovered these two methods of telomere maintenance may occur concurrently (21, 22).

We analysed how POLD3 depletion may affect telomere length in CRC cell lines. As seen in Figure 4A and B, both CACO2 shRNA1 and shRNA2 and RKO shRNA1 and shRNA2 cells showed a significant decrease in telomere length after 15 days of POLD3 knockdown (Both CACO2 and RKO: P<0.05, 1 μg/mlDox and P<0.01, 10 μg/ml Dox). Decreased telomere length varied with increased POLD3 depletion, suggesting a direct link between POLD3 levels and telomere length.

**Figure 4:**
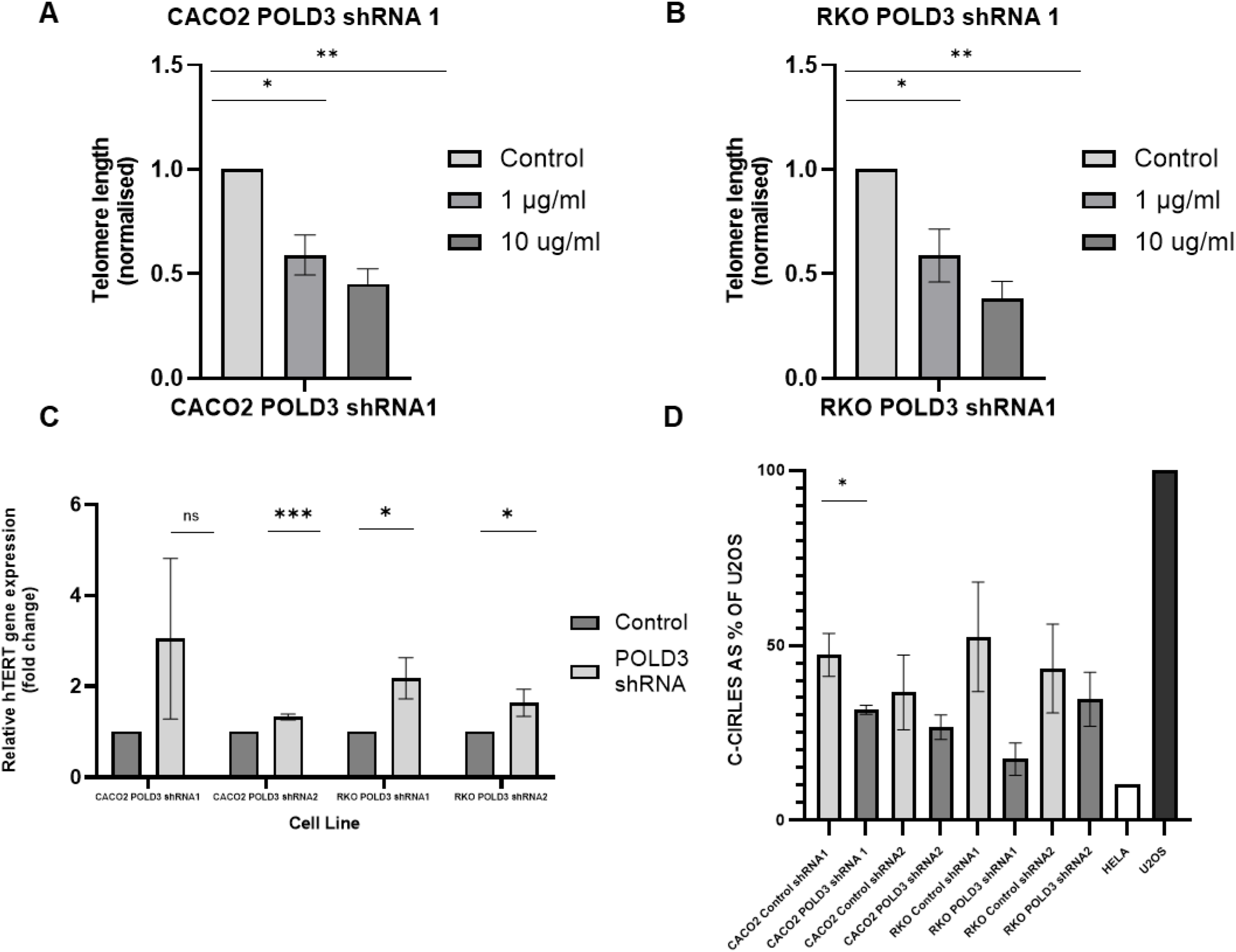
POLD3 knockdown results in shortened telomeres and disruption of the ALT pathway A) Telomere length analysis of CACO2 POLD3 shRNA1 cells treated with 0 (DMSO), 1 μg/ml or 10 μg/ml doxycycline to induce knockdown of POLD3. Doxycycline and control treatments were refreshed every 72hrs for 15 days. Control with 1 μg/ml P<0.05 and 10 μg/ml P<0.01. (T-test, N=3). B) Telomere length analysis of RKO POLD3 shRNA1 cells treated with 0 (DMSO), 1 μg/ml or 10 μg/ml doxycycline to induce knockdown of POLD3. Doxycycline and control treatments were refreshed every 72hrs for 15 days. Control with 1 μg/ml P<0.05 and 10 μg/ml P<0.01. (T-test N=3). C) qPCR analysis of hTERT mRNA expression in CACO2 POLD3 shRNA1, CACO2 POLD3 shRNA, RKO POLD3 shRNA1 and RKO POLD3 shRNA2 cells treated with 0 (DMSO), or 10μg/ml doxycycline to induce shRNA expression and POLD3 knockdown for 14 days. CACO2 POLD3 shRNA2 P<0.003, RKO POLD3 shRNA1 P< 0.029, RKO POLD3 shRNA2 P<0.004. D) C-circle analysis of CACO2 POLD3 shRNA1, CACO2 POLD3 shRNA, RKO POLD3 shRNA1 and RKO POLD3 shRNA2 cells treated with 0 (DMSO), or 10μg/ml Doxycycline to induce shRNA expression and POLD3 knockdown after 7 days incubation. CACO2 shRNA 1: P<0.0477 (T-test). One way ANOVA across all cell lines: P<0.035. N=4. Error bars given as ± SEM in all panels.

Initially, TERT levels in our POLD3 knockdown cell lines were tested. Unexpectedly we found an increase in TERT expression compared to our control cells (Figure 5 C) (CaCO2 shRNA1 NS, CaCO2 shRNA2 P<0.001, RKO shRNA1 P<0.05, RKO shRNA2 P<0.05) despite the shortened telomeres. It is possible that this increase in TERT does not result in telomerase activity and may also be a response to the shortening telomeres such as been previously observed in normal human cells (23). In contrast, when we assessed the ALT pathway we did see a reduction in activity, in line with our observations of telomere length. Figure 5 D shows an observable decrease in the presence of c-circles, a known hallmark specific to the ALT mechanism, in POLD3 knockdown cell lines. C-circles were presented as a % of c-circle’s present in cell line U2OS, which is known to harbour high ALT activity (24). The reduction was significant in CACO2 cells with shRNA 1 (P<0.05) with other cell lines displaying the same trend. One way ANOVA across all cell lines revealed a significant reduction in the presence of C-circles (P<0.05). Taken together these data suggest that a decrease in POLD3 reduces the activity of the ALT pathway of telomere maintenance and that hTERT expression increases in response to shortened telomeres, possibly as a compensatory mechanism.

## Discussion

POLD3, part of the Polδ complex is important for DNA synthesis during cellular proliferation, DNA damage repair via the BIR pathway, and also telomere maintenance via the ALT pathway.

Our analysis of the TCGA COAD dataset suggested that lower levels of POLD3 expression are predictive of reduced overall survival, with the strongest association in MSS cancers. Given the essential role of POLD3 in replication fidelity and repair of replication-associated lesions (13, 25), reduced expression of its canonical isoform may predispose to replication stress, particularly in MSS tumours where mismatch repair remains intact.

Our functional studies were therefore designed to test whether modest reductions in POLD3 are sufficient to perturb cancer cell behaviour and genome maintenance. POLD3 knockdown in CRC cell lines reduced cell proliferation as expected due to its role in DNA replication, with cells partially stalling at S-phase. However, we suggest that its role in DNA repair is more critical for cancer progression with knockdown cells exhibiting increased DNA damage and DSB accumulation. Over time a modest reduction in POLD3, would increase levels of DNA damage, and associated genome instability. As an extension of this we have shown that POLD3 knockdown reduces telomere length, potentially creating further instability. Together these findings suggest an important role for POLD3 in growth and maintenance of cancer cells.

We have clearly shown that POLD3 loss results in impaired proliferation of CRC cells, evidenced by slower growth over time, reduced colony forming capacity, reduction of Ki67+ cells, and stalling of cell cycle progression. The intrinsic need for cancer cells to proliferate to form tumours relies of DNA polymerases, including POLD3, suggesting that reduction of POLD3 may result in slower growing tumour phenotypes. Indeed, complete knockout of POLD3 in mice has been shown to cause embryonic lethality, suggest POLD3 is essential for DNA replication (26). POLD3 mRNA expression was reduced to approximately half of the control expression in our cell lines. These POLD3-depleted cell numbers were reduced to half to two thirds of that of the control cells after 96 hours with the reduction in mRNA levels reflecting the reduction in growth as well as increased apoptosis We hypothesise that further knockdown of POLD3 could not be achieved in these cancer cells and would result in lethality.

Alongside this decrease in growth which could be interpreted to suggest that POLD3 reduction has a tumour suppressive effect, we also observed an increase in DNA damage. We demonstrated both increased comet tail length and an accumulation of γH2AX foci indicating a general increase in double strand breakage. This increased damage was accompanied by upregulation of common DNA repair pathway proteins, ATM and RAD21, which are essential in repair of DSBs by HR. ATM binds to and aids in the phosphorylation of γH2AX, thus serving as a central transducer to recruit further repair machinery, such as MRN (27). Damaged DNA is excised and repair is initiated by RAD21, before new DNA synthesis is mediated by POLD3 (28). Interestingly the increase in both ATM and RAD21 expression, suggesting activation of DNA repair pathways involving HR, is unable to diminish the numbers of γH2AX foci. POLD3 depletion may reduce the efficacy of DNA damage repair, despite upregulation of repair proteins earlier in the pathway.

Next, we analysed the effects of POLD3 depletion on telomere length. Telomeres are usually maintained by the Telomerase pathway, mediated by the catalytic TERT protein, which is activated in 90% of tumours. Alternatively, telomeres can be maintained by the ALT pathway, which utilises BIR, in which POLD3 is essential. Our cell lines have previously been shown to be TERT positive, with relatively high levels of TERT found in CACO2 and RKO cells (29, 30).

Telomere length was approximately half of that of the control upon POLD3 depletion after 15 days consecutive treatment. This was despite increased TERT expression, which may have been caused by activation of the TERT gene in response to short telomere recognition (23). We therefore suggest that telomere shortening due to POLD3 depletion is due to disruption of the ALT pathway. In support of this, we have shown the presence of C-circles in our cell lines, a hallmark of the ALT pathway. C-circles in our control cells were roughly 50% of that in U2OS cells (known for their high ALT activity), and far higher than in the ALT negative cell line HELA, which showed less than 10% of C-circles present in U2OS. Upon POLD3 depletion, the levels of C-circles present were reduced by nearly half in some cell lines, suggesting a direct reduction in ALT mechanism upon POLD3 depletion.

Previous data has suggested that the ALT pathway is only activated in response to TERT negative status, or in response to TERT abolishment (31, 32). However, we have shown both activation of the *TERT* gene, and the presence of C-circles, in our cell lines, suggesting in these cells that ALT occurs concurrently with telomerase-mediated maintenance. Therefore, upon ALT reduction due to POLD3 loss, cells may become reliant on telomerase activation. Alternatively, POLD3 depletion may also cause DSBs at telomere ends during replication, in which case TERT is activated, but cannot keep up with the DNA damage inflicted, resulting in shortened telomeres.

Together, these data present an interesting role of POLD3 in the context of CRC. POLD3 depletion causes decreased proliferation and stalling of cell cycle progression, which is often a positive target for the treatment of CRC. However, with POLD3 depletion we also see a manifestation of DNA instability, shown by increased presence of DSBs, and activation of DNA repair pathways. Furthermore, we have shown that POLD3 depletion results in shortened telomeres, which may create further genome instability in our cell lines, but also may reduce the longevity of cancer cells. It appears that at this level of knockdown, over time the levels of genome instability and reduction in proliferation combine to drive the cells to apoptosis.

Taken together, the existing genetic data, and our functional data point to an important, context-dependent role for POLD3 in CRC. If POLD3 is indeed the target of CRC risk SNPs in 11q13.4, we hypothesize that small reductions in POLD3 expression, in intestinal cells that are already proliferating fast and/or have pre-cancerous mutations, may increase genetic instability and accrual of DNA damage without substantial reductions in proliferation. This is particularly evident in cancers which lack other key drivers of genetic instability (eg. MMR or other polymerase proofreading mutations).

In conclusion, we propose that the 11q13.4 risk SNPs may reduce effective POLD3 dosage in colorectal cancers, thereby promoting increased DNA damage and telomere shortening, and providing a plausible mechanistic link between germline genetic risk and tumour genomic instability.

## Methods

### Survival Analysis

TCGA colon adenocarcinoma (COAD) RNA-seq and clinical data were analysed using TCGAbiolinks. Harmonized GDC STAR-count data were retrieved for primary tumours and normal colon tissues, and one tumour sample per patient was retained. Counts were normalized with edgeR and transformed to log-counts per million (logCPM). POLD3 expression was examined according to microsatellite status (MS), consensus molecular subtype (CMS) and pathological stage. Microsatellite annotations were obtained from TCGA subtype information, and CMS labels were assigned using CMScaller. Differences in POLD3 expression across subgroups were assessed using one-way ANOVA and Kruskal-Wallis tests, with post hoc pairwise Wilcoxon rank-sum tests corrected by the Benjamini-Hochberg method. Overall survival analyses were restricted to tumours with available follow-up time and event data, with death coded as the event and patients alive at last follow-up treated as censored observations. Kaplan-Meier curves were generated after median-based stratification of POLD3 expression and compared using the log-rank test. Associations between continuous POLD3 expression and overall survival were assessed using Cox proportional hazards models, with multivariable analyses adjusted for age, sex and pathological stage.

### Cell culture and maintenance

Immortalised human colorectal adenocarcinoma cell lines CACO2 and RKO (acquired from ATCC) were maintained in Gibco Dulbecco’s Modified Eagle Medium (DMEM) (Sigma-Aldrich) supplemented with 10% foetal bovine serum (FBS) (Sigma-Aldrich), and 1% penicillin streptomycin (Sigma-Aldrich). Cells were grown in an humified atmosphere at 37◦c with 5% CO2. Subculturing was performed every 72 hours to maintain a cell confluency of < 80%.

### Generation and validation of POLD3 shRNA

POLD3 shRNA target sequences were obtained from Broad institute GPP web portal (Clone ID: TRCN0000233258, TRCN0000233261) and were inserted into shRNA sequences as described here. shRNAs were cloned into EZ-Tet-pLKO-Puro (Addgene #85966) followed by validation by Sanger sequencing and restriction digest. The vector was then transfected in HEK293 cells along with viral packaging vectors (2^nd^ generation system – pCMV-dR8.2 and pCMV-VSV-G, Addgene #8455 and #8454) using Lipofectamine 2000 (Invitrogen, US). Virus containing media was collected, sterilized and titre measured (Go-Stix, Takara). The cell lines CACO2 and RKO were transduced, and cells with integrated EZ-Tet-pLKO-Puro-POLD3 were selected for with puromycin. To confirm overexpression, doxycycline was added at low (0.1 μg/ml), medium (1 μg/ml) or high (10 μg/ml) levels. RNA was extracted (RNeasy, QIAGEN) and quantified by real-time reverse transcriptase polymerase chain reaction (qPCR) using TaqMan technology according to the manufacturers protocol (Applied biosystems). Each assay was repeated in triplicate. Relative expression values were calculated using the ΔΔCt method using GAPDH normalisation.

### shRNA targets

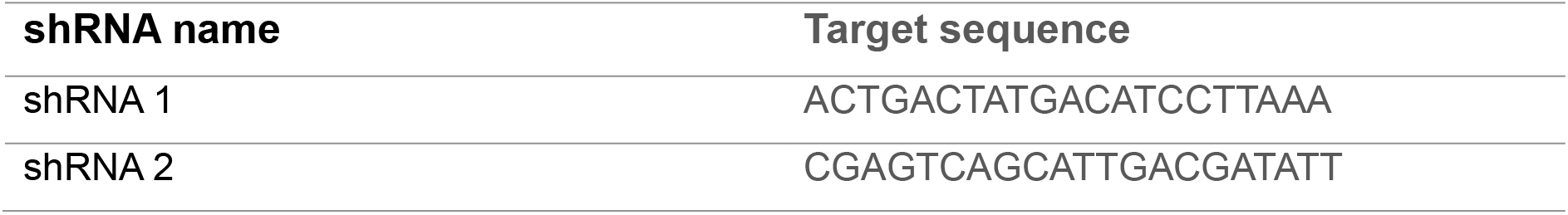

### TaqMan probes

| **Gene name** | **Probe ID** |
|---|---|
| POLD3 | Hs01075255 |
| GAPDH | Hs99999905_m1 |

### Western blot

For intracellular protein detection, cells were lysed by resuspension in RIPA buffer. 30 μg of protein were loaded per sample. Protein samples were separated via 4-12% sodium dodecyl sulfate polyacrylamide gel electrophoresis under denaturing conditions, and then transferred onto the nitrocellulose membrane (Millipore, UK) under 20 V. Membranes were blocked with 5% milk for 1hr at room temperature. Membranes were then incubated with primary antibody in TBST-5% BSA overnight at 4◦C. Membranes were then washed with TBST. Secondary antibody was added for 1 hr at room temperature. Membranes were imaged through incubation with Enhanced chemiluminescence (ECL). The ratio of optical density of the bands was measured by a gel image analysis system (Bio-Rad) and normalized to B-actin as a loading control.

### Antibodies used

|  |
|---|
| POLD3 (1:1000. Catalogue number:). |
| Goat anti rabbit (1:5000. Catalogue number:). |

### Cell proliferation assay

To assess cellular proliferation during POLD3 knockdown, cells were plated at a density of 5×10^3^ in replicate with DMSO control, low (0.1 μg/ml), medium (1 μg/ml) or high (10 μg/ml) doxycyclinetreatments. Cellular proliferation was assessed via MTT (sigma-Merck) according to the manufacturers instruction and the absorbance read at 560/640 nm at at 24, 48 and 72 hrs. The results were analysed using GraphPad Prism.

### Immunofluorescence

For cellular protein detection, cells were plated on coverslips and grown to ∼70% confluency. Cells were fixed with methanol, and the cellular membrane was permeabilised with TRITON X 0.5% for 5 minutes. Cells were blocked with 1% BSA for 1 hr at 37◦C, and then incubated with primary antibodies in 1% BSA for 1hr at 37◦C. A secondary antibody was then added to cells for 1hrs at 37◦C. 5 μl of mounting media with DAPI (VECTASHIELD® WZ-93952-27, Cole-Parmer, UK) was then placed onto the coverslips, and coverslips were fixed on to slides for imaging.

### Antibodies used

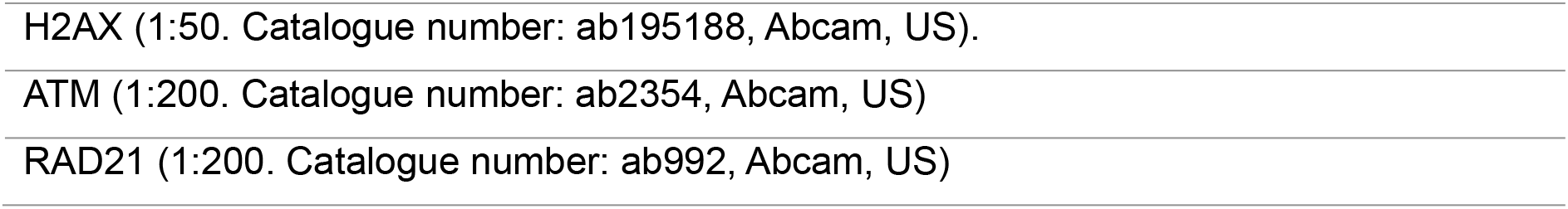
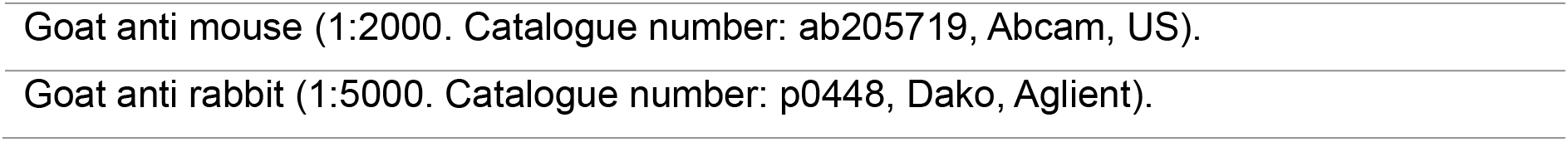

|  |
|---|
| H2AX (1:50. Catalogue number: ab195188, Abcam, US). |
| ATM (1:200. Catalogue number: ab2354, Abcam, US) |
| RAD21 (1:200. Catalogue number: ab992, Abcam, US) |
| Goat anti mouse (1:2000. Catalogue number: ab205719, Abcam, US). |
| Goat anti rabbit (1:5000. Catalogue number: p0448, Dako, Aglient). |

### Comet assay

Cells were plated at a density of 1×10^5^ in with DMSO control, low (0.1 μg/ml), medium (1 μg/ml) or high (10 μg/ml) levels doxycycline treatment. After 72hrs, cells were harvested and diluted in PBs and a density of 10^4^. Cell suspension was mixed 1:5 with low-gelling agarose and 100 μl was placed on Polylysine coated slides dipped in 1% agarose (Sigma-Aldrich) and allowed to set. A further 100 μl low gelling agarose (Sigma-Aldrich) was placed on top and allowed or set. Cell lysis was then performed by submerging slides in lysis buffer (2% N-Lauroylsarcosine sodium salt (Sigma-Aldrich), 0.5M NA_2_EDTA (Sigma-Aldrich) and 0.1mg/ml proteinase K) for 1hr. Slides were then washed in Electrophoresis buffer for 1.5 hrs (90 mM Tris Buffer (Fisher Bioreagents), 90 mM boric acid (Sigma-Aldrich) and 2 mM NA_2_EDTA). Slides were then placed in an electrophoresis tank and submerged in electrophoresis buffer, under 20V current for 40 mins. Slides were then stained with 1% SYBR SAFE (Invitrogen) in TBE for 20 mins, before dehydration through submersion in 70%, 90% and 100% ethanol. Slides were visualised by Leica DM4000.

### Telomere length analysis

To assess telomere length, real-time PCR was used. Relative telomere length was compared to that of a single copy gene (36B4) to control for amplification of each sample, and to determine genome copies per sample. Genomic DNA was extracted from our POLD3 knockdown cell lines (DNeasy, QIAGEN) treated with doxycycline at 0.1 μg/ml, 1 μg/ml, and 10 μg/ml or DMSO. Telomere length analysis was performed as described. Briefly, two q-PCR master mixes were prepared with either the copy gene primer (36B4) or the telomere primer. Master mixes prepared as followed: 10 μl 2X Power SYBER Green PCR Master Mix (Applied Biosystems), 2 μM forward primer (Telomere-F or 36B4-f), 2 μM reverse primer (Telomere-R or B6B4-R), 4 μl of 5ng/μl DNA, and nuclease free water to 20 μl. Each sample was prepared in triplicate. Primers were used as described below.

A standard curve was generated using serial dilutions of 36B4 standard (6125000 kb to 6.125 kb). Plasmid DNA (pBR322) was also added to each standard to maintain a constant 20ng of total DNA per reaction tube. A telomere standard curve was established by serial dilution of the telomere standard (1018400 kb to 10184 kb) and was used to measure content of telomeric sequence per sample. Real-time PCR runs were performed in triplicate for each of the DNA pools.

### Primers

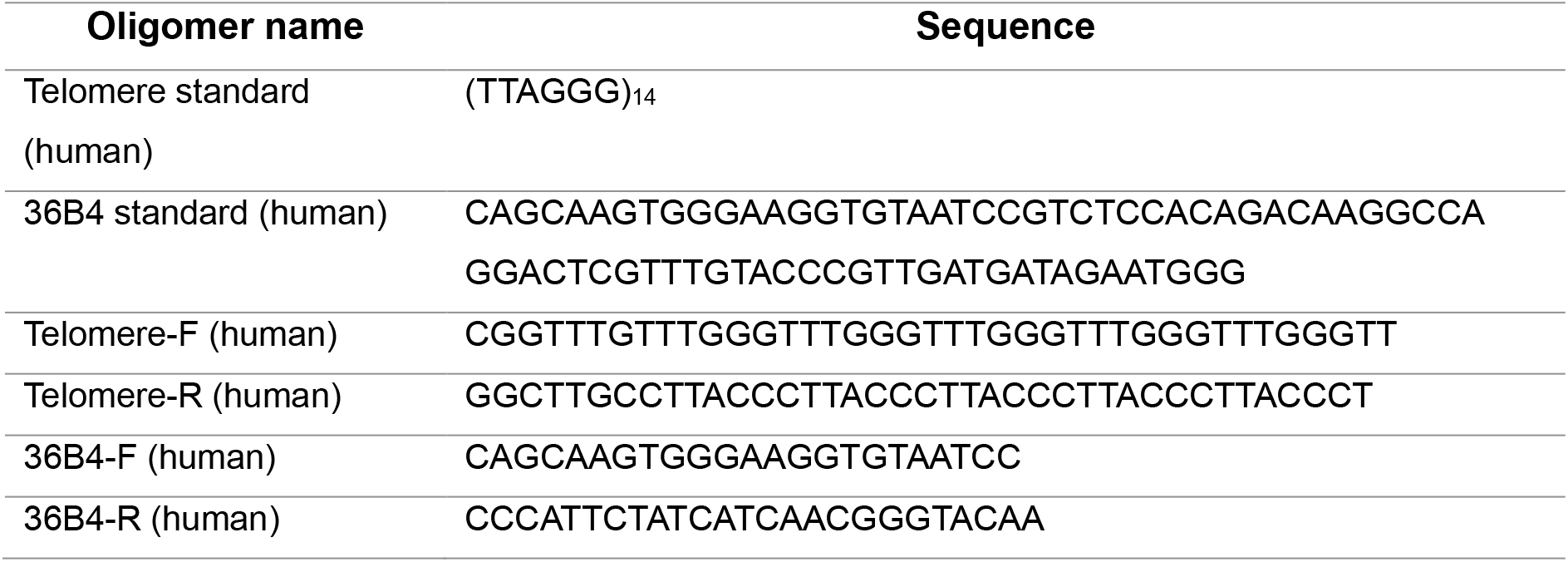

### C-circle analysis

To assess the level of c-circle present, real time PCR was used. C-circle levels were determined as a % of total c-circles present in cell line U2OS. The method for C-circle amplification was a previously described.

Briefly, genomic DNA was extracted from our POLD3 knockdown cell lines (DNeasy, QIAGEN) treated with doxycycline at 10 μg/ml, or DMSO. Rolling cycle of C-circle amplification was performed using 40 ng total genomic DNA and made up to 10 μl with 10 mM Tris. To diluted DNA 10 μL of master mix was added: 0.8 μl 10 M DTT, 2 μl Phi 29 DNA pol buffer (NEB, UK), 0.4 μl 10 mg/ml BSA, 0.2 μl 10% tween, 0.8 μl 1 mM dTTP, 1.5 μl phi 29 DNA pol (NEB), 4.3 μl water. Separate samples were also made without phi 29 DNA pol as a control. Samples were run for 8 hrs at 30 °C and deactivated at 65°Cfor 20 minutes. qPCR on amplified C-circle DNA was then performed as described before in telomere length analysis.

## Data Availability

All data produced in the present study are available upon reasonable request to the authors

## Acknowledgements

We thank E Efendi for help with image quantification and cell culture maintenance.

## Author Contributions

A.L., E.C., and N.D.L conceived the study. N.D.L carried out expression and survival analyses. E.C and A.L. carried out cell line generation and growth and DNA damage analyses. E.C., S.A. and T.R. carried out telomere length and pathway analysis. A.L., E.C and N.D.L wrote the manuscript. All authors critically reviewed the manuscript content. All authors read and approved the final manuscript.

