## Supplementary Figures for "Reduced POLD3 Expression Induces DNA Damage and Telomere Shortening in Colorectal Cancer Cells: A Potential Mediator of Colorectal Cancer Risk Variants"

### Supplementary Figure S1

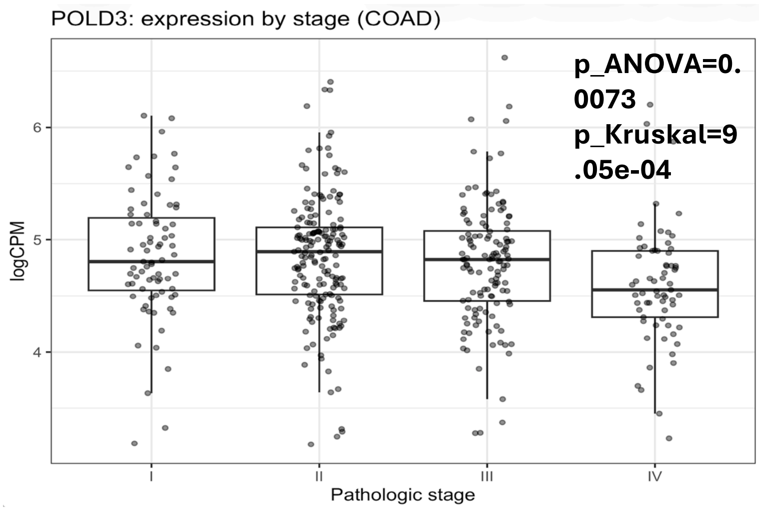


**Supplementary Figure S1. POLD3 expression by pathological stage in TCGA colon adenocarcinoma (COAD).** POLD3 expression across pathological stages I-IV in the TCGA COAD cohort. Expression values are shown as logCPM from RNA-seq data. Boxplots represent the median and interquartile range, with individual tumours overlaid. Global differences across stages were assessed using one-way ANOVA and Kruskal-Wallis tests, with post hoc pairwise Wilcoxon rank-sum tests corrected by the Benjamini-Hochberg method (Supp Table S1).

### Supplementary Figure S2

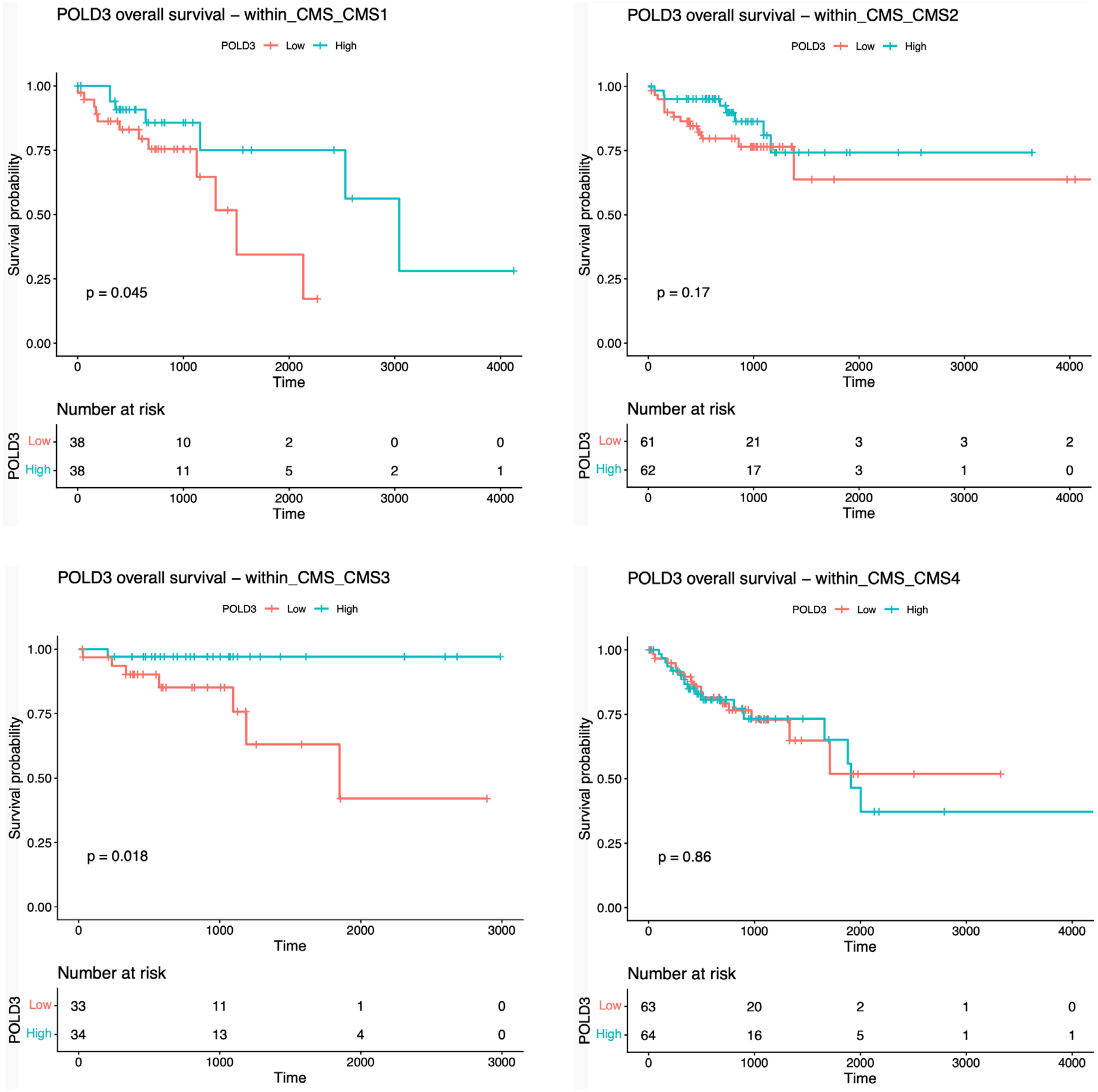


**Supplementary Figure S2. CMS-stratified Kaplan–Meier survival analyses according to POLD3 expression in TCGA COAD.** Kaplan-Meier analyses of overall survival in TCGA COAD stratified by median POLD3 expression within individual CMS subtypes (CMS1-CMS4). In each panel, tumours were divided into low and high POLD3 expression groups based on the within-subtype median. Tick marks indicate censored observations. Full subgroup-specific Kaplan-Meier and Cox results are provided in the Supplementary Tables S2-S3.

### Supplementary Figure S3

A

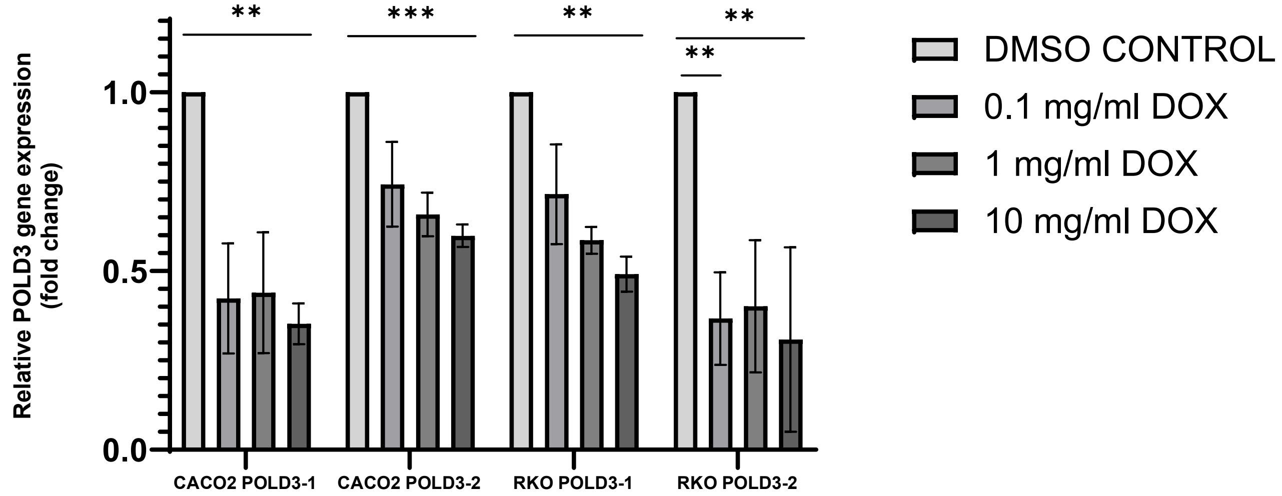


B

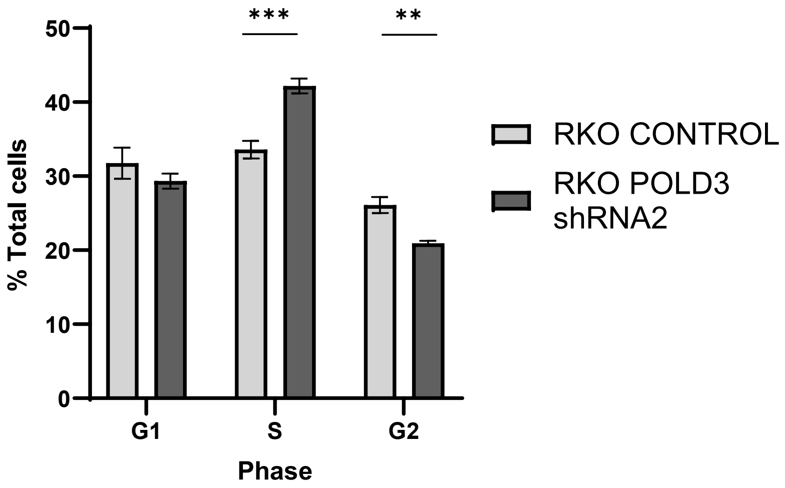


**Supplementary Figure S3** A) POLD3 mRNA levels in cell lines with lentiviral transduction of shRNA-1 and shRNA-2 POLD3 knockdown constructs as measured by Q-PCR using taqman technology. Cells were treated with 0 (DMSO only), 0.1, 1 or 10 μg/ml Doxycyline for 48 hours to induce shRNA expression and POLD3 knockdown. B Quantification of cell cycle stage of RKO POLD3 shRNA 2 induced with 10 μg/ml doxycyline. Student T-test: shRNA 2: S $P<0.005$, G2<0.001. Error bars given as ± SEM in all panels.
